# The validation of a mobile sensor-based neurobehavioral assessment with machine learning analytics

**DOI:** 10.1101/2021.04.29.21256265

**Authors:** Kelly L. Sloane, Shenly Glenn, Joel A. Mefford, Zilong Zhao, Man Xu, Guifeng Zhou, Rachel Mace, Amy E Wright, Argye E. Hillis

**Author notes:** Corresponding Author: Shenly Glenn, 260 King St., Unit 1505, San Francisco, CA 94107, USA, Text. No authors are US government employees. Author Contributions: Conceptualization: SG, JAM, AEH Methodology: JAM, SG, AEH Software: MX, ZZ, GZ Validation: GZ, MX, ZZ Formal analysis: JAM, ZZ, SG, GZ Investigation: KLS, RM, AEW, AEH Data curation: GZ Draft preparation: KLS, SG, JAM Manuscript Review: MX, ZZ, GZ, RM, AEW, AEH.

## Abstract

**Background:** Mild cognitive impairment is a common yet complex condition that is often underdiagnosed in the early stages. Miro Health is a mobile platform for the self-administration of sensor-based cognitive and behavioral assessments that was developed to measure factors typical of legacy neuropsychological tests in addition to behaviors that are currently left to subjective clinical impression such as eye movement, language, and processing speed.

**Objective:** Studies were conducted to measure Miro Health’s concurrent validity, test-retest reliability, and amnestic MCI classification performance.

**Method:** Spearman correlations were calculated to estimate the concurrent validity of Miro Health variables with legacy neuropsychological test variables using data from **160** study participants. Fifty-nine healthy controls were assessed at three time points to evaluate the test-retest reliability of Miro Health scores. Reliability was quantified with the scores’ intraclass correlations. Learning effects were measured as trends. In addition, a machine learning algorithm combined Miro Health variable scores into a Risk Score designed to distinguish 65 healthy controls (HC), 38 MCI participants (21 amnestic MCI (aMCI), and 17 non-amnestic MCI (naMCI)).

**Results:** Significant correlations of Miro Health variables with legacy neuropsychological test variables were observed. Longitudinal studies show agreement of subsequent measurements and minimal learning effects. The Risk Score distinguished aMCI from healthy controls with an Area Under the Receiver Operator Curve (AUROC) of 0.97; the naMCI participants and controls were separated with an AUROC of 0.80, and the combined MCI group (aMCI + naMCI) was separated from healthy controls with an AUROC of 0.89.

**Conclusion:** Miro Health includes valid and reliable versions of variable scores that are analogous to legacy neuropsychological variable scores and a machine-learning derived risk score that effectively distinguishes HCs and individuals with MCI.

## Introduction

Mild Cognitive Impairment (MCI) is a complex clinical disorder with many possible causes including neurodegenerative diseases, traumatic head injury, stroke or other vascular disorders, cancer, medication effect, or other medical issues. It is estimated that 20-40% of MCI cases will progress to dementia (Roberts et al., 2013), while other cases of MCI may remain static or resolve. For progressive cases, the early identification of MCI and timely support of patients and their care communities may help prolong independence, reduce healthcare costs and advance research toward effective therapies (Lee et al., 2020; Wittenberg et al, 2019).

To aid in early identification and continual monitoring, diagnostic tools are required that can be administered repeatedly and reliably and can detect subtle deviations from healthy cognition and behavioral function (Sabbagh et al., 2020). Cognitive assessments performed by licensed clinical neuropsychologists remain the gold standard for the diagnosis of MCI (Petersen et al., 2018). Yet, clinician-administered assessments are impractical for population-based care due to their limited availability and expense. Brief screens like the Montreal Cognitive Assessment and the Mini Mental Status Exam can be rapidly administered and interpreted without neuropsychology training (Folstein et al., 1975; Nasreddine et al., 2005; Seo et al., 2011), but their ability to detect early MCI varies depending on patient population and is limited by practice effect. Gaze trackers have emerged as a potential rapid assessment but lack complementary cognitive measures and require the purchase of external hardware, thus limiting their accessibility (Oyama et al., 2019). Their diagnostic utility and reliability remain unknown.

This paper presents validity studies for a new mobile assessment platform that combines cognitive and neurobehavioral measures for the detection and characterization of MCI called ‘Miro Health’. This platform is a mobile application that offers self-administered assessments in the form of interactive modules and self-report questionnaires. Two versions of Miro Health are validated in this paper: V.2 and V.3. Improvements made to V.2 tutorials, stimuli display times, and guided prompts changed the characteristics of the data collected, and thus necessitated a new mobile version (V.3), an updated V.3-matched Reference Data Set, and separate V.3 validity analyses. There were three main aims of this study. The first aim was to demonstrate validity of Miro V.2 and V.3 by measuring the correlation of variable scores common to legacy neuropsychological tests and Miro. The second aim was to evaluate the stability of Miro’s measurement properties by test-retest reliability. Additionally, the third aim investigates the predictive validity of a machine-generated “aMCI Risk Score” to distinguish healthy performance from amnestic MCI (aMCI), and aMCI from non-amnestic MCI (naMCI). Studies were overseen by an external study monitor.

## Methods

A total of 209 participants were recruited for this study from neurology clinics at Johns Hopkins and the general public. Participant sample is summarized in Table 2. HCs were recruited through ads in newspapers. Screening measures included: demographics; medical history (self-reported); Telephone Interview for Cognitive Status (TICS); Geriatric Depression Scale; and the Mini Mental Status Exam (MMSE). *HC inclusion criteria*: Ages 65 and over; score of 33 or greater on the TICS; English speaker prior to age 5; high school or equivalent education. *Affected inclusion criteria*: Ages 65 and over; score of 14-26 on the MMSE; diagnosis of MCI or neurodegenerative disorder; vascular disorder with cognitive impairment. *Exclusion criteria*: Evidence of comorbid neurological disease, use of drug known to affect cognition; uncorrected vision or hearing impairment; history of cancer, substance abuse, or axis 1 disorder.

This study was approved by the Johns Hopkins University Institutional Review Board (protocol 00088299) and New England IRB (protocols 120180208, 120180211, 120180209 and 12080253) for the use of human subjects. Capacity was determined and consent was obtained from each participant.

### Computerized Cognitive Assessment

The Miro Health platform features mobile cognitive and behavioral assessments that can be self-administered. The mobile application can be downloaded to mobile devices from iTunes or Google Play. It takes 5-60 minutes to self-administer, depending on the assessment battery, with automated scoring. Miro has been designated as a Breakthrough Device by the FDA. The Miro Health assessment batteries may be tailored from a comprehensive library of self-report questionnaires and more than 40 interactive modules. Many Miro modules are redesigned analogs of legacy neuropsychological tests (Table 1) that have been updated to capture high-fidelity data like movement, speech, language, and timing through sensors built-in to their mobile devices, (e.g. touchscreen, microphone), in order to quantify behavioral functions in addition to cognitive functions. Each Miro module is automatically and consistently administered, adaptive to patient performance, and equipped with patented and proprietary anti-cheat mechanisms (Glenn et al, 2017; Glenn and Mefford 2019b).

**Table 1.** Variable examples

| **Legacy test**<br>Administered in-person by a licensed clinical neuropsychologist | **Miro module**<br>Self-Administered, mobile device | **Miro variable examples** |
| --- | --- | --- |
| Iowa Trail Making Task (A and B) | Bolt Bot | Total Time B – Total Time A |
| Simple Reaction Time, No comparator | Bosco | Response time (post-correct – correct) |
| Category Fluency (D-KEFS) | Categories | Total correct (phonemic – category) |
| Phonemic Fluency (D-KEFS) | Lucky Letters | Slope correct responses, 60s |
| Design Fluency A (D-KEFS) | Chart A Course | Linear optimality |
| Stroop Test (PAR) | Colors and Words | Avg response time (Colors – Words) |
| Picture memory, No comparator | Deja Vu | Total (correct – errors) / total time |
| No equivalent, No comparator | Family Farm | Total correct |
| Spatial Span Forward & Backward (WMS) | Follow the Glow | Avg inter-response interval |
| Digit Span Forward (WAIS-IV) | Hungry Bees | Longest correct span |
| Digit Span Backward (WAIS-IV) | Hungry Bees | Longest correct span |
| Sentence Repetition (V2.0 National Alzheimer's coordinating Center) | Repeat After Me | Word order |
| Hopkins Verbal Learning Test (HVLT-R) Immediate Recall | Spy Games | Avg response latency |
| HVLT-R, Delayed Free Recall | Spy Report | Total correct – Total errors |
| HVLT-R, Delayed Cued Recall | Spy Report | Total errors, commission |
| Picture Description | Speak the Scene | Number of content units, right |
| Picture Description | Speak the Scene | Fundamental frequency |
| Finger Tapping Test | Take Flight | Avg taps, dominant hand |
| Clock Drawing | Tick Tock | Number correct |
| Design Fluency B (D-KEFS) (Switching) | Treasure Hunt | Avg inter-response interval (B – A) |
| Coding (WAIS-IV) | Treasure Tomb | Slope errors, 60s |
| Choice Reaction Time, No equivalent | Two of A Kind | Avg response time (complex – simple) |
| Confrontation Naming (BNT-15) | Wordy Goat | Response time, standard deviation |

*Data processing* includes proprietary digital signal processing (A.I.) that extracts features from recordings, distills variables from those features, and prepares distilled variables for analysis (Glenn et al, 2019a). In addition to familiar variables like out-of-set and repeat errors, commission and omission errors, and total test time, hundreds of other categorical and continuous variables are automatically extracted and calculated (see Table 1 for variable examples).

*Data analysis* includes automated machine learning algorithms (A.I.) that identify relevant variables, combine and weight those variables, and then compare results to Reference Data Sets to predict symptom identification, symptom severity, and diagnosis.

### Statistical Analyses

All of the statistical analyses were performed using R software, R 4.0.3 (www.R-project.org).

#### Concurrent Validity

A total of 160 participants were included in the concurrent validity study; 67 patients were recruited from neurology clinics and 93 healthy controls (HCs) from the general public; 28 participants were excluded for incomplete data sets (22 incomplete legacy neuropsychological data sets, 6 incomplete Miro data sets). Half of the participants received Miro first, half received legacy tests first, alternating by even and odd enrollee numbers. All participants were tested on the same Miro mobile battery; unique Miro module versions were presented at each assessment with the exception of Picture Description, Category Fluency, and Letter Fluency. Legacy test administration was conducted with two separately ordered test batteries that consisted of the same assessments. The battery was alternated upon odd number enrollee triggers, e.g., enrollee 1 and 2 received the battery order A, enrollee 3 and 4 received battery order B, enrollee 5 and 6 received battery order A, and so on.

Participants were excluded from the correlation of individual variables when missing either Miro or comparator scores (or both) for that variable. Standardization of scores occurred via a two-step process: (1) Scores were quantile normalized or had quantiles in the empirical distribution of scores mapped to quantiles of a standard normal distribution; (2) The mean (normalized) score for normal participants was subtracted from each score and the resulting centered scores were rescaled by the standard deviation of the scores among the normal participants. This process saw that standardized scores for healthy controls have mean values of zero and standard deviations of one and an overall normal distribution. For scores derived from Category Fluency and Letter Fluency modules, scores based on test sessions with different stimuli were standardized separately.

Spearman correlations were used to analyze concurrent validity between individual variable scores produced by Miro’s automated data capture and scoring approach to the analogous neuropsychological test scores administered and scored by licensed clinical neuropsychologists (Signorell et al, 2020). For comparison, Pearson correlations were calculated as well and are shown with the Spearman correlations in the supplement.

### Test-Retest Reliability

A total of 65 healthy participants were recruited from the general public; 6 participants were excluded from the study for failing Miro Health’s proprietary anti-cheat mechanism built into the application. Thirty-three participants completed the test-retest reliability study using Miro V.2 and 26 using Miro V.3 (see Table 2). Half of the participants were unsupervised, half were supervised, alternating by even and odd enrollee numbers.

**Table 2.** Participant demographics

|  | Group | Participants | Female / Male | Avg Age<br>(range) in yrs |
| --- | --- | --- | --- | --- |
| Concurrent Validity V.2 | Healthy Control | 19 | 15 / 4 | 73.2 (64 – 89) |
|  | Affected | 27 | 16 / 11 | 78.5 (68 – 95) |
| Concurrent Validity V.3 | Healthy Control | 74 | 41 / 33 | 70.1 (61 – 82) |
|  | Affected | 40 | 11 / 29 | 69.3 (48 – 83) |
| Test-retest V.2 | Healthy Control | 33 | 25 / 8 | 74.2 (65 – 81) |
| Test-retest V.3 | Healthy Control | 26 | 16 / 10 | 72.0 (64 – 82) |
| Predictive Validity V.3 | Healthy Control | 65 | 36 / 29 | 70.2 (61 – 82) |
|  | aMCI | 21 | 6 / 15 | 71.1 (49 – 83) |
|  | naMCI | 17 | 6 / 11 | 71.1 (60 – 82) |

MCI=mild cognitive impairment, aMCI = amnesic MCI, naMCI = non-amnesic MCI

Enrolled participants were evaluated with the Miro platform over three assessments at one-week intervals. The study battery occurred in the same order at each time point. A different version of each module was randomly generated for each participant at each time point with the exception of Picture Description, Letter Fluency, and Category Fluency whose versions were presented in the same order.

The test-retest reliability of scores generated over 3 time points was quantified by estimating the intra-class correlation (ICC), an estimate of the variance in the distribution of scores that can be explained by individual participant random effects in a mixed effect model (Gamer, 2019). In addition to the reliability of measurements, learning effects or trends across the three assessments were analyzed using two measures named ‘trends’ and ‘progression’. Estimated ‘trends’ were calculated for each variable score as the slope of a linear model for score versus observation number. The ‘progression’ was calculated as the difference in slopes between the first and second assessments and between the second and third assessments. The statistical significance of trends and progressions were tested with Wald-tests.

### Predictive Validity

A total of 139 participants were included in the predictive validity study; 103 V.3 participants (38 MCI and 65 HC) and 36 V.2 participants (17 MCI and 19 HC). Data collected from the concurrent validity study and time point one of the test-retest reliability study were included in the predictive validity study based on the following criteria: (1) HC: must have more than 85% of the data set present and be ages 65 and over; (2) MCI: must meet American Academy of Neurology clinical criteria for MCI diagnosis (American Academy of Neurology, 2018), MMSE score cannot be lower than 20 (Chapman et al, 2016; O’Bryant 2008).

Predictive validity group assignments were made by a licensed clinical neuropsychologist who had access to participant scores on legacy neuropsychological tests (Table 1), the Mayo-Portland Modified Inventory, the geriatric depression scale, as well as demographics and medical history. Of the 103 V.3 participants, the neuropsychologist identified 65 healthy controls and 38 with MCI and assigned MCI participants to an amnestic MCI (aMCI) sub-type when impaired performance in the domain of learning and memory was evident or to a non-amnestic MCI (naMCI) sub-type when impaired performance in 1 or more non-memory domains was evident. MCI sub-type analyses were not conducted for V.2 participants due to low total MCI numbers.

Machine learning approaches were used to find an optimal weighted combination of variable scores useful in the assignment of study participants to their expert-designated group. Inputs to the model came from variables derived from the battery of administered Miro modules (Table 1), including variables available from legacy neuropsychological tests and others not captured from traditional testing modalities such as continuous timing measures, and the quantification of movement, vocal characteristics, speech production, and language (Table 1). Variables were quantile normalized and missing values were imputed (Hastie et al, 2013). The regression algorithm in the OrdinalNet package v.2.9 (Wurm et al, 2017) was used to fit an ordinal logistic regression model with elastic net penalties and to identify a subset of variables most relevant for classification and find a weighted combination of the selected scores that best separates the aMCI, naMCI, and control groups.

The regression parameters, alpha (the ratio of L1 to L2 penalties) and lambda (the scaling factor for the penalties), were optimized for classification performance as measured by AUROC with 5-fold cross-validation. Classification models were not adjusted for age, sex, or education due to the small sample size and risk of overfitting the model. The resulting weighted combination of input scores will be referred to as the “aMCI Risk Score”.

The algorithm was evaluated by leave-one-out cross-validation. The resulting aMCI Risk Score formula was applied to the left-out individual’s data set to get an out-of-sample predicted risk score. This was done for each observation in turn and the AUROC was calculated using the set of out-of-sample risk scores as a classifier to separate the groups. For comparison, individual variable AUROCs for group separation were also calculated. For data collected with Miro V.2, MCI Risk Scores were generated using the same approach of elastic net (Zou et al, 2005) but with only two groups, HC and MCI, whereas three groups were analyzed in the Miro V.3 analysis.

## Results

### Concurrent Validity

All participants were assessed with comparator neuropsychological tests; 46 were also assessed with Miro V.2 (27 affected, 19 healthy control) and 114 with Miro V.3 (40 affected, 74 healthy control). Table 3 shows the comparator tests and paired Miro modules considered for the concurrent validity analysis. Correlations of scores from Miro V.2 (Table S1) and Miro V.3 (Table 4) show concurrent validities as estimated by Spearman correlations ranging from 0.36 to 0.60 and 0.27 and 0.68, respectively. Table S2 includes results of both Pearson and Spearman correlations for the concurrent validity analysis of Miro V.3 and demonstrates that the two results are strongly in agreement. Data for V.3 concurrent validity can be found in the supplemental file, V.3_CV.xlsx.

**Table 3.**
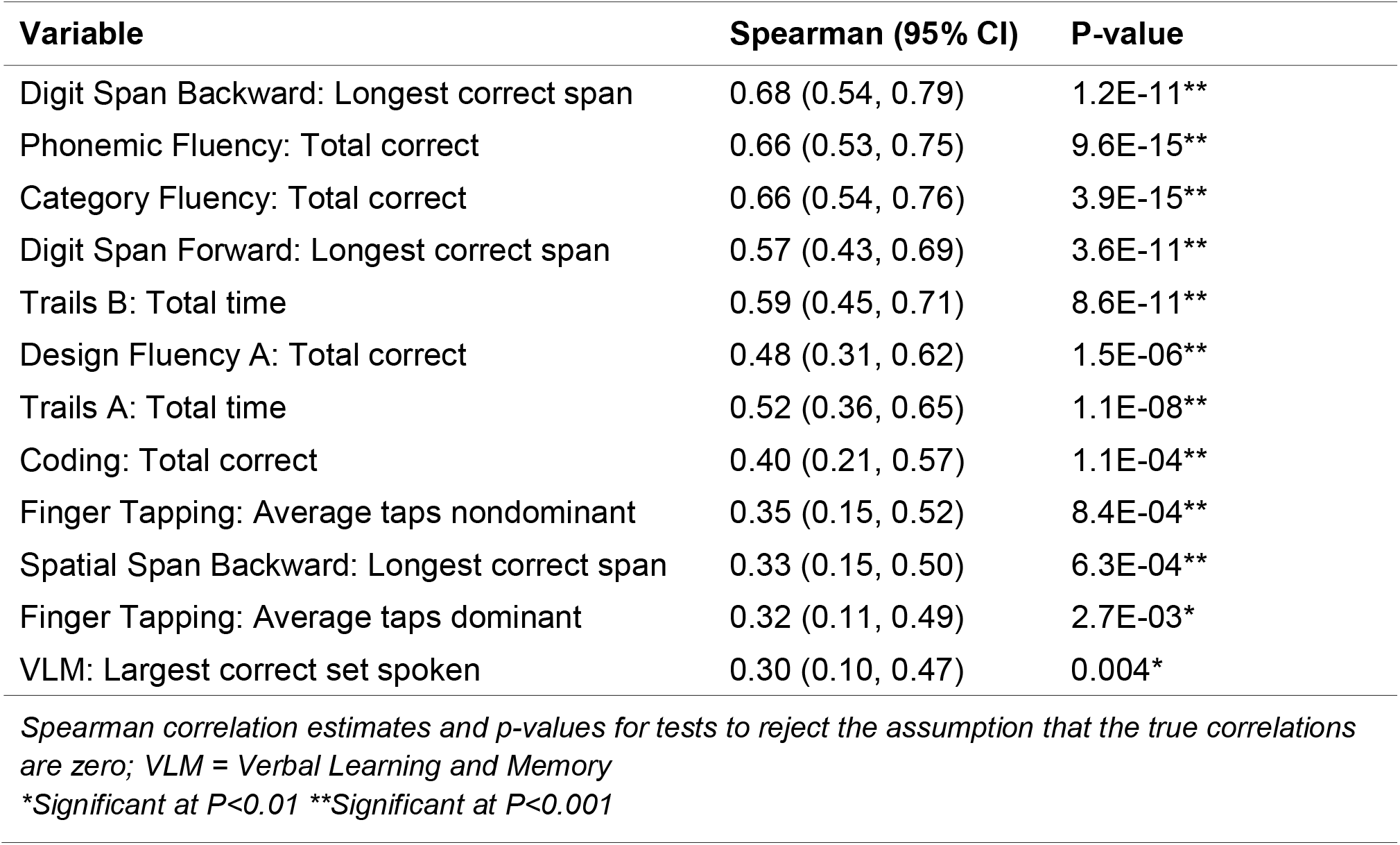
Concurrent validity of Miro Version 3 vs. comparative neurocognitive tests

| Variable | Spearman (95% CI) | P-value |
| --- | --- | --- |
| Digit Span Backward: Longest correct span | 0.68 (0.54, 0.79) | 1.2E-11** |
| Phonemic Fluency: Total correct | 0.66 (0.53, 0.75) | 9.6E-15** |
| Category Fluency: Total correct | 0.66 (0.54, 0.76) | 3.9E-15** |
| Digit Span Forward: Longest correct span | 0.57 (0.43, 0.69) | 3.6E-11** |
| Trails B: Total time | 0.59 (0.45, 0.71) | 8.6E-11** |
| Design Fluency A: Total correct | 0.48 (0.31, 0.62) | 1.5E-06** |
| Trails A: Total time | 0.52 (0.36, 0.65) | 1.1E-08** |
| Coding: Total correct | 0.40 (0.21, 0.57) | 1.1E-04** |
| Finger Tapping: Average taps nondominant | 0.35 (0.15, 0.52) | 8.4E-04** |
| Spatial Span Backward: Longest correct span | 0.33 (0.15, 0.50) | 6.3E-04** |
| Finger Tapping: Average taps dominant | 0.32 (0.11, 0.49) | 2.7E-03* |
| VLM: Largest correct set spoken | 0.30 (0.10, 0.47) | 0.004* |

*Spearman correlation estimates and p-values for tests to reject the assumption that the true correlations are zero; VLM = Verbal Learning and Memory*
*\*Significant at $P < 0.01$ \*\*Significant at $P < 0.001$*

**Table 4.** Intraclass correlations for Miro versions of legacy scores

| Variable | ICC (95% CI) |
| --- | --- |
| Design Fluency Switching: Total correct | 0.79 (0.65, 0.89)* |
| Trails B: Total time | 0.76 (0.62, 0.86)* |
| Phonemic Fluency: Total correct | 0.75 (0.61, 0.85)* |
| Trails A: Total time | 0.74 (0.60, 0.85)* |
| DVLM: Total correct | 0.72 (0.47, 0.86)* |
| Coding: Total correct | 0.61 (0.42, 0.76)* |
| Digit Span Backward: Longest correct span | 0.58 (0.23, 0.78)* |
| Digit Span Forward: Longest correct span | 0.58 (0.23, 0.78)* |
| VLM: Largest correct set spoken | 0.56 (0.34, 0.74)* |
| DVLM: Total errors | 0.54 (0.19, 0.76)* |
| Design Fluency A: Total correct | 0.53 (0.33, 0.70)* |
| Finger Tapping: Average taps dominant | 0.42 (0.21, 0.63)* |
| Spatial Span Forward: Total correct | 0.40 (0.19, 0.61)* |

*VLM = Verbal Learning and Memory; DVLM = Delayed Verbal Learning and Memory;*
*\*Significantly different than zero at the 0.05 level*

### Test-Retest Reliability

Table 5 shows Miro V.3 variables with legacy correlates have ICC estimates between 0.40 and 0.79 Table S3 shows the estimated trends and progression for these variables. No variable demonstrated significant trends or progressions. For reference, collected Miro V.3 study data for concurrent validity and test-retest reliability are included in the supplemental materials.

**Table 5.** Performance (AUROC) of the MCI risk score and ten legacy test scores on separation of healthy controls from MCI participants using diagnoses from either a Neuropsychologist or a brief screen

| Score | aMCI<br>vs HC | naMCI<br>vs HC | MCI vs<br>HC | aMCI vs<br>naMCI |
| --- | --- | --- | --- | --- |
| Trails A: Total time | 0.94 | 0.69 | 0.83 | 0.84 |
| Category Fluency: Total correct | 0.92 | 0.69 | 0.82 | 0.72 |
| Coding: Total correct | 0.91 | 0.68 | 0.80 | 0.75 |
| Trails B: Total correct | 0.89 | 0.69 | 0.80 | 0.75 |
| Phonemic Fluency: Total correct | 0.88 | 0.79 | 0.84 | 0.67 |
| Trails B: Total time | 0.85 | 0.72 | 0.80 | 0.70 |
| Digit Span Forward: Longest correct span | 0.85 | 0.55 | 0.70 | 0.83 |
| Picture Description: Total prepositions | 0.84 | 0.76 | 0.76 | 0.75 |
| Digit Span Backward: Longest correct span | 0.82 | 0.57 | 0.70 | 0.81 |
| Design Fluency: Total correct | 0.81 | 0.63 | 0.72 | 0.69 |
| MCI Risk score | 0.97 | 0.80 | 0.89 | 0.83 |

### aMCI Predictive Validity

In the V.3 data set, when healthy control, aMCI, and naMCI labels are assigned by brief screen scores, Miro’s aMCI risk score yields a classification performance measured as an AUROC of 0.83. When a licensed neuropsychologist assigns labels, Miro’s aMCI risk score has an AUROC of 0.97 for classification of aMCI vs. HC and an AUROC of 0.89 for the classification of MCI (aMCI + naMCI) vs. HC. The V.2 MCI Risk Score classified the MCI and HC participants with an AUROC of 0.92.

For comparison, individual variable AUROCs for group separation were also calculated. Results are shown in Table 6. In Table S4, the means and standard deviations of non-normalized scores in the HC and aMCI groups are shown, along with p-values from Wilcoxon rank-sum tests and t-tests.

## Discussion

In this paper we report the results of three validity studies that examine Miro Health’s ability to characterize cognitive function. There were three specific aims of this study: (1) to demonstrate the correlation of familiar neuropsychological test variables with analogous Miro Health variables so that the capture and extraction of fundamental units of data which serve as inputs into Miro Health’s machine learning engine could be validated; (2) to demonstrate the test-retest reliability of Miro’s modules; and (3) to investigate the capabilities of an automated self-administered, machine-learning system to identify aMCI performance scores and to differentiate aMCI scores from those of naMCI and HC scores.

Results show that the Miro Health mobile, self-administered platform automatically classifies the performance of HCs and those with MCI as well as current standards for participants within this data set. This study further demonstrates substantial differences in AUROCs when labels are assigned by brief screen scores (AUROC 0.83) vs. a licensed clinical neuropsychologist (AUROC 0.97), a consequence of the poor measurement properties of brief screens. The modest concurrent validity correlations evidenced in this study were expected given the modest test-retest reliability of comparator tests (Iverson, 2001; Snow et al, 1989).

Unlike typical test-retest validity studies that analyze data collected across two time points, this study analyzed data collected across three time points. This design was chosen in order to examine the effects that increasing familiarity with mobile interfaces and assessment paradigms may exhibit on performance scores. Familiarity effects were evaluated by testing for trends or average changes in scores on successive assessments and by testing for progression, or differences, in the changes in performance between the 1^st^ and 2^nd^ assessments and between the 2^nd^ and 3^rd^ assessments. Neither type of familiarity effect was significant for the scores evaluated in this study. This suggests that Miro Health’s unlimited, equivalent module versions resist learning effects.

The strong performance of Miro’s machine-learning derived classifier is the result of a combination of factors that are brought together in the Miro Health Platform, including that: (1) In addition to cognitive data, behavioral data like movement, speech, language, and voice are quantified and incorporated into the classifier. (2) Self-administered assessments and machine-driven scoring removes the variability, errors, and subjectivity found in human administration and scoring of cognitive assessments. (3) Miro Health’s built-in Quality Management System ensures data validity at all stages of assessment, processing and scoring. For example, Miro Health’s Transcription Platform acts like an ‘Uber for Transcription’. It alerts human transcribers as soon as new data hits the backend. The first two respondents independently transcribe the data. The differences between the two human transcriptions and the machine transcription are automatically highlighted. A third human reviewer is then alerted to make a final determination so that variables can be extracted and prepared for scoring. (4) Machine learning approaches interrogate data to find informative features and optimally combine them to make a classifier suited to the purpose of distinguishing particular groups of study participants and their source populations.

Many important topics raised by Miro Health’s novel approach warrant investigation. This paper did not present analyses of supervised vs. unsupervised assessment results, the effectiveness of Miro Health’s anti-cheat detection, the capture and creation of new variable types, the value of categorical vs. continuous data in MCI detection, or the relative merits of specific modules for their ability to detect and sub-type MCI. A benefit of Miro Health’s modular platform is that it can rapidly deploy and scale investigations into questions like these, accelerating the advancement of cognitive and behavioral research and rapidly incorporating relevant research results into healthcare delivery.

Study limitations include: (1) small samples sizes; (2) the lack of diverse diseases and symptom severities; and (3) the lack of longitudinal data. Larger sample sizes would allow more stable estimates of the weights for combined scores and the evaluation of score performance as a classifier by cross-validation and hold-out sets. Data from diverse diseases and symptom severities would help demonstrate Miro Health’s real-world ability to identify aMCI against other similar conditions such as cancer-related cognitive impairment or cognitive impairment due to anti-coagulant use. Longitudinal data collection would allow retrospective relabeling of data once symptoms had progressed and the accuracy of diagnosing MCI’s underlying pathology improved. Longitudinal data would also facilitate a move away from the use of threshold scores for the diagnosis of MCI and toward more personalized measures of change over time.

## Conclusion

A machine-learning derived risk score based on mobile, self-administered cognitive and behavioral tests distinguished aMCI participants from HCs with an Area Under the Receiver Operator Curve (AUROC) of 0.97, while the naMCI participants and controls were separated with an AUROC of 0.80, and the combined MCI group (aMCI + naMCI) was separated from HCs with an AUROC of 0.89.

The data demonstrates that a self-administered mobile cognitive and behavioral assessment combined with an automated A.I. scoring platform differentiates MCI from HC as well as current methods in two sequential validity studies and analyses, Miro V.2 and Miro V.3. Miro Health’s cost-effective approach supports massively parallel data collection and analyses to improve healthcare, accelerate research, and reduce costs.

## Data Availability

Data used in the paper is temporarily not available for public.

## Acknowledgment of any presentation of the material

None

## Supplement S1. Miro Health Platform Description

Miro Health was founded with the mission of making quality comprehensive brain healthcare accessible and affordable to everyone. The Miro Health platform brings modern data science techniques to the measurement of human cognition and behavior. Miro has been in development and testing since 2013 and has been designated as a Breakthrough Device by the FDA. Miro mobile applications were designed in parallel with extensive usability studies on patients with varying cognitive and physical abilities. Miro Health’s platform consists of interactive, gamified brain assessments that can be accessed from the convenience of a mobile device (phone or tablet) by downloaded from the iTunes or Google Play stores. The Miro Health assessment batteries may be tailored from a comprehensive library of self-report questionnaires and more than 40 interactive modules. Many Miro modules are redesigned analogs of legacy neuropsychological tests that have been updated to capture high-fidelity data like movement, speech, language, and timing through sensors built-in to mobile devices, (e.g. touchscreen, microphone). Each Miro module is automatically and consistently administered, adaptive to patient performance, and equipped with patented and proprietary anti-cheat mechanisms. Depending on the number of modules selected, the assessment can take 5 to 60 minutes.

The platform consists of four primary components: administrative management; data capture; data processing; and data analysis.

*Administrative management* automates many of the administrative tasks of clinical research, healthcare delivery, and patient management.

*Data capture* offers mobile, self-administrable assessments for download to iOS and Android phones and tablets. Miro Health assessment batteries may be tailored from a comprehensive library of self-report questionnaires and more than 40 interactive modules. Many Miro modules are redesigned analogs of legacy neuropsychological tests (Table 3) that have been updated to capture high-fidelity data like movement, speech, language, and timing in order to quantify behavioral functions in addition to cognitive functions. While clinically informative, these data have not been routinely collected as a first-step in the clinical care pathway due to the impracticality of the multiple specialists required as well as the time and expense involved. Each Miro module is automatically and consistently administered, adaptive to patient performance, and equipped with patented and proprietary anti-cheat mechanisms (Glenn et al, 2017; Glenn and Mefford 2019b). As patients interact with Miro modules, sensors built-in to their mobile devices, (e.g. touchscreen, microphone), capture high fidelity, unmediated data for analysis. Each user session triggers a unique presentation of the same Miro module while maintaining parameter consistency and equivalent measurements (Glenn and Mefford, 2020). This patented approach was designed to provide unlimited versions of the same assessment to facilitate reliable, repeated data capture.

*Data processing* includes proprietary digital signal processing (A.I.) that extracts features from recordings, distills variables from those features, and prepares distilled variables for analysis (Glenn et al, 2019a). In addition to familiar variables like out-of-set and repeat errors, commission and omission errors, and total test time, hundreds of other categorical and continuous variables are automatically extracted and calculated from recorded patient interactions (see Table 1 for variable examples).

Miro incorporates and derives *machine-learning* classifiers. Machine learning approaches interrogate data to find informative features and optimally combine them to make a classifier suited to the purpose of distinguishing particular groups of study participants and their source populations. Self-administered assessments and machine-driven scoring removes the variability, errors, and subjectivity found in human administration and scoring of cognitive assessments.

Miro Health’s built-in *Quality Management System* ensures data validity at all stages of assessment, processing and scoring. For example, Miro Health’s Transcription Platform acts like an ‘Uber for Transcription’. It alerts human transcribers as soon as new data hits the backend. The first two respondents independently transcribe the data. The differences between the two human transcriptions and the machine transcription are automatically highlighted. A third human reviewer is then alerted to make a final determination so that results can be compared to automated speech-to-text performance and variables can be extracted and prepared for scoring.

**Table S1.** Concurrent validity of Miro V.2 vs. comparison neurocognitive tests

| Score | Spearman (95% CI) | P-value |
| --- | --- | --- |
| Digit Span Backward: Longest correct span | 0.60 (0.37, 0.76) | 1.5E-05*** |
| Coding: Total correct | 0.58 (0.34, 0.75) | 3.1E-05*** |
| Design Fluency A: Total correct | 0.58 (0.34, 0.75) | 4.3E-05*** |
| Digit Span Forward: Longest correct span | 0.49 (0.23, 0.68) | 5.4E-04*** |
| Category Fluency: Total correct | 0.49 (0.23, 0.68) | 5.9E-04*** |
| Finger Tapping: Avg taps dominant | 0.50 (0.21, 0.71) | 1.5E-03** |
| Phonemic Fluency: Total correct | 0.42 (0.14, 0.63) | 3.9E-03** |
| Finger Tapping: Avg taps nondominant | 0.36 (0.04, 0.61) | 0.027* |

\*Significant at $P < 0.05$ \*\*Significant at $P < 0.01$ \*\*\*Significant at $P < 0.001$

**Table S2.** Comparison of Spearman and Pearson correlations for concurrent validity

| Score | Spearman<br>(95% CI) | P-value | Pearson<br>(95% CI) | P-value |
| --- | --- | --- | --- | --- |
| Phonemic Fluency: Total correct | 0.66 (0.53, 0.75) | 9.6E-15<sup>***</sup> | 0.66 (0.54, 0.76) | 5.3E-15<sup>***</sup> |
| Category Fluency: Total correct | 0.66 (0.54, 0.76) | 3.9E-15<sup>***</sup> | 0.59 (0.46, 0.70) | 1.0E-11<sup>***</sup> |
| Digits Backward: Longest correct | 0.68 (0.54, 0.79) | 1.2E-11<sup>***</sup> | 0.70 (0.56, 0.80) | 1.9E-12<sup>***</sup> |
| Digits Forward: Longest correct | 0.57 (0.43, 0.69) | 3.6E-11<sup>***</sup> | 0.62 (0.49, 0.72) | 2.3E-13<sup>***</sup> |
| Trails B: Total time | 0.59 (0.45, 0.71) | 8.6E-11<sup>***</sup> | 0.52 (0.36, 0.65) | 3.0E-08<sup>***</sup> |
| Trails A: Total time | 0.52 (0.37, 0.65) | 1.1E-08<sup>***</sup> | 0.43 (0.26, 0.58) | 4.9E-06<sup>***</sup> |
| Coding: Total correct | 0.40 (0.21, 0.57) | 1.1E-04<sup>***</sup> | 0.40 (0.21, 0.56) | 1.1E-04<sup>***</sup> |
| Spatial Span Back: Total correct | 0.33 (0.15, 0.50) | 6.3E-04<sup>***</sup> | 0.41 (0.23, 0.56) | 2.4E-05<sup>***</sup> |
| Finger Taps: Avg nondominant | 0.35 (0.15, 0.52) | 8.4E-04<sup>***</sup> | 0.26 (0.06, 0.44) | 0.013* |
| Finger Taps: Avg dominant | 0.32 (0.11, 0.49) | 2.7E-03<sup>**</sup> | 0.27 (0.06, 0.45) | 0.011* |
| VLM: Largest correct set spoken | 0.30 (0.10, 0.47) | 0.004* | 0.27 (0.06, 0.45) | 0.010* |
| Design Fluency B: Total correct | 0.28 (-0.02, 0.54) | 0.064 | 0.18 (-0.13, 0.46) | 0.250 |
| Phonemic Fluency: Error set loss | -0.10 (-0.28, 0.09) | 0.302 | -0.08 (-0.26, 0.11) | 0.409 |
| Phonemic Fluency: Error repeat | 0.06 (-0.13, 0.24) | 0.561 | 0.28 (0.10, 0.44) | 3.2E-03<sup>**</sup> |
| Category Fluency: Error repeat | 0.04 (-0.15, 0.23) | 0.668 | -0.04 (-0.22, 0.15) | 0.717 |
| Design Fluency B: Total errors | -0.08 (-0.37, 0.22) | 0.594 | -0.14 (-0.42, 0.17) | 0.383 |
| Category Fluency: Error set loss | -0.15 (-0.33, 0.04) | 0.130 | -0.05 (-0.24, 0.13) | 0.573 |
| Design Fluency A: Total errors | 0.01 (-0.24, 0.22) | 0.936 | -0.06 (-0.26, 0.15) | 0.604 |

VLM = Verbal Learning and Memory; DVLM = Delayed Verbal Learning & Memory
\*Significant at $P < 0.05$ \*\*Significant at $P < 0.01$ \*\*\*Significant at $P < 0.001$

**Table S3.** Test-retest analysis of learning effects

| **Score** | **Trend**<br>**Avg (SD)** | **Trend**<br>**p-value** | **Progression**<br>**Avg (SD)** | **Progression**<br>**p-value** |
| --- | --- | --- | --- | --- |
| Category Fluency: Total correct | -0.019 (0.55) | 0.99 | -0.12 (1.8) | 0.99 |
| Category Fluency: Error set loss | 0.000 (0.50) | 1.00 | 0.35 (1.9) | 0.97 |
| Coding: Total correct | -0.089 (0.40) | 0.97 | 0.27 (1.6) | 0.97 |
| Design Fluency A: Total correct | 0.100 (0.51) | 0.97 | -0.25 (1.4) | 0.97 |
| Design Fluency Switch: Error repeat | 0.025 (0.72) | 0.99 | 0.37 (1.7) | 0.97 |
| Design Fluency A: Total errors | -0.130 (0.57) | 0.96 | -0.10 (1.9) | 0.99 |
| Design Fluency Switch: Total correct | 0.043 (0.31) | 0.98 | -0.13 (1.1) | 0.98 |
| Design Fluency Switch: Error repeat | -0.063 (0.39) | 0.97 | -0.55 (1.7) | 0.95 |
| Finger Tap: Avg taps nondominant | 0.002 (0.47) | 1.00 | -0.031 (1.1) | 1.00 |
| Finger Tap: Average taps dominant | -0.025 (0.46) | 0.99 | -0.43 (1.2) | 0.95 |
| Phonemic Fluency: Total correct | -0.060 (0.30) | 0.97 | -0.29 (1.1) | 0.96 |
| Phonemic Fluency: Error repetitions | 0.063 (0.50) | 0.98 | 0.26 (1.6) | 0.97 |
| Trails A: Total time | -0.053 (0.32) | 0.97 | 0.54 (1.2) | 0.93 |
| Trails B: Total time | -0.080 (0.32) | 0.96 | 0.20 (1.1) | 0.97 |
| VLM: Largest correct set spoken | 0.036 (0.51) | 0.99 | 0.39 (1.3) | 0.95 |
| VLM: Error set loss | -0.035 (0.44) | 0.99 | -0.06 (1.1) | 0.99 |
| VLM: Error repeat | -0.048 (0.27) | 0.97 | -0.22 (2.4) | 0.99 |

**Table S4.** Means, standard deviations, and tests of differences in distribution of Miro scores in Healthy Control and aMCI subjects

| Score | HC<br>Avg (SD) | aMCI<br>Avg (SD) | Rank sum<br>p-value | t-test<br>p-value |
| --- | --- | --- | --- | --- |
| Trails A: Total time | 16 (4.0) | 28 (8.7) | 1.6E-09<sup>***</sup> | 8.9E-13<sup>***</sup> |
| Category Fluency: Total correct | 13 (5.0) | 4.1 (3.7) | 7.5E-09<sup>***</sup> | 6.8E-11<sup>***</sup> |
| Phonemic Fluency: Total correct | 17 (5.0) | 8.3 (5.4) | 2.0E-07<sup>***</sup> | 1.3E-09<sup>***</sup> |
| Trails B: Total time | 87 (21) | 110 (14) | 1.3E-06<sup>***</sup> | 2.7E-06<sup>***</sup> |
| VLM: Error set loss | 0.05 (0.21) | 0.62 (1.0) | 4.7E-04<sup>***</sup> | 1.4E-04<sup>***</sup> |
| VLM: Total errors | 1.1 (2.6) | 2.3 (4.1) | 1.1E-03<sup>**</sup> | 0.19 |
| VLM: Error repeat | 1.1 (2.6) | 1.7 (3.5) | 1.8E-03<sup>**</sup> | 0.46 |
| Phonemic Fluency: Error repetitions | 1.8 (1.7) | 1.1 (1.6) | 0.05 | 0.12 |
| Design Fluency A: Total correct | 14 (3.8) | 9.4 (3.7) | 0.07 | 8.6E-05<sup>***</sup> |
| Phonemic Fluency: Error set loss | 0.67 (1.7) | 1.2 (1.5) | 0.09 | 0.18 |
| Design Fluency A: Error set loss | 2.8 (2.5) | 2.5 (1.8) | 0.33 | 0.65 |
| Category Fluency: Error repeat | 0.67 (1.0) | 0.52 (1.0) | 0.34 | 0.58 |
| Design Fluency A: Total errors | 5.1 (2.8) | 4.8 (3.1) | 0.37 | 0.72 |
| Coding: Total correct | 37 (11) | 21 (8.2) | 0.41 | 4.7E-06<sup>***</sup> |
| VLM: Largest correct set spoken | 7.5 (5.0) | 4.9 (1.5) | 0.66 | 0.07 |
| Category Fluency: Error set loss | 1.7 (3.5) | 1.9 (2.6) | 0.67 | 0.88 |

VLM = Verbal Learning and Memory; DVLM = Delayed Verbal Learning and Memory
\*Significant at $P < 0.05$ \*\*Significant at $P < 0.01$ \*\*\*Significant at $P < 0.001$

